# E-field guided repetitive transcranial magnetic stimulation modulates oscillatory brain activity dynamics in tinnitus

**DOI:** 10.1101/2025.11.18.25340332

**Authors:** Stefan Schoisswohl, Berthold Langguth, Patrick Neff, Martin Schecklmann, Tobias Kleinjung, Payam S. Shabestari

## Abstract

**Introduction:** The auditory phantom sound perception tinnitus is accompanied by maladaptive neurophysiological changes. In tinnitus treatment, repetitive transcranial magnetic stimulation (rTMS) is applied to counteract these pathological alterations. Previous work showed that single-session rTMS can reduce tinnitus loudness and modulate tinnitus-associated oscillatory brain activity. This study aimed to contribute to this research branch by addressing previous methodological shortcomings, including the absence of neuronavigation and adequate sham control. The objective was to assess tinnitus loudness and ongoing brain activity changes following various rTMS protocols, and to characterize modulations related to patient-specific most effective protocols, potentially uncovering electrophysiological markers of tinnitus suppression.

**Methods:** Three active protocols (1,10,20 Hz; 200 pulses) and one sham protocol (0.1 Hz; 20 pulses) were delivered to the left and right temporo-parietal junction of 22 chronic subjective tinnitus patients using e-field-guided neuronavigation. Resting state EEG was recorded before and after each stimulation, along with tinnitus loudness ratings. Patient-specific protocols eliciting maximal suppression were identified via significant pre-to-post loudness reductions exceeding sham.

**Results:** Right 10 Hz rTMS induced strongest loudness suppressions. Significant and sham-superior EEG modulations were observed after right 10 Hz, right 20 Hz and left 20 Hz. Power increases in the Delta, Theta, Alpha and Gamma frequency bands were mainly observed in frontal and temporal areas but did not correlate with reported tinnitus suppression. In 16 patients it was feasible to identify a protocol inducing significant loudness reduction exceeding sham. Here suppression was associated with increased Alpha activity in the frontal cortex.

**Conclusions:** Our findings demonstrate that brief rTMS protocols can transiently suppress tinnitus and modulate tinnitus-related oscillatory brain activity dynamics. Frontal Alpha power increases might reflect local enhanced inhibitory processes and reduced tinnitus percept processing, emphasizing frontal Alpha as a candidate marker for effective tinnitus suppression.

## I. INTRODUCTION

Repetitive transcranial magnetic stimulation (rTMS) is a safe and well-established non-invasive neuromodulation technique applied in basic brain research and in the treatment of several neuropsychiatric conditions [1–4]. Combinations with neurophysiological measures such as electroencephalography (EEG) have demonstrated its ability to modulate evoked and spontaneous brain activity dynamics, with effects outlasting the stimulation offset. Hence, neuroplastic consequences can be induced [5–11]. It is assumed that low frequency rTMS (≤ 1 Hz) inhibits, and high frequency rTMS (≥ 5 Hz) excites neural activity [3,4,12,13]. However, this heuristic is only a rough guide, as many investigations report contradictory findings or variability in effects between and within subjects [14–17].

Chronic subjective tinnitus is defined as the persistent and conscious auditory perception of tonal or composite noise without any corresponding peripheral acoustic source [18]. Tinnitus is thought to emerge from diminished peripheral acoustic input due to noise trauma or hearing loss [19], which results in maladaptive neuroplastic alterations such as hyperactivity in central auditory-related structures or abnormal neural synchrony and bursting along the auditory pathway [20–23]. On a macroscopic level these pathological alterations are reflected in tinnitus-associated neural oscillations such as increased activity in the Delta, Theta and Gamma frequency ranges or decreased Alpha band activity in auditory brain areas as demonstrated by various investigations using EEG or magnetoencephalography (MEG) [24–30]. These abnormalities in spontaneous brain activity were subsumed under the frameworks of the Thalamo-Cortical Dysrhythmia model (TCD) and the Synchronization-by-Loss-of-Inhibition-Model (SLIM) [26,31,32].

Traditionally, 1 Hz rTMS over the left auditory cortex is applied in the treatment of tinnitus with the rational to suppress tinnitus-associated hyperactivity and thus reduce tinnitus loudness and suffering [33,34]. Despite recent reports of efficacy [35,36], overall findings remain inconsistent and are determined by inter-individual variability in treatment responses and inconclusive meta-analytic/review evidence [1,37–40]. In experimental studies using different kinds of stimulation protocols, it was demonstrated that single rTMS sessions are able to elicit transient suppressions of the tinnitus percept as well [41–45].

Combining such single sessions with measures of spontaneous brain activity not only enables evaluations of immediate neuroplastic rTMS effects or links between M/EEG activity and tinnitus suppression but also facilitates deeper insights into the pathophysiology behind tinnitus perception.

To the best of our knowledge, up to now two studies were conducted in this regard. In an MEG study, Müller et al. tested various tonic and patterned magnetic stimulation protocols over the left auditory cortex and found an increase in auditory cortex Alpha band power (8-12 Hz) linked to tinnitus loudness reduction [46]. Schecklmann et al. could demonstrate in an EEG study that 1 Hz rTMS applied to the left temporal cortex induced power decreases in the Delta (2-3.5 Hz) and Theta (4-7.5 Hz) frequency bands as well as an increase in Beta2 power (18.5-21 Hz) in prefrontal sensors. A right-hemispheric frontal cortex 1 Hz stimulation resulted in a reduction of Beta3 (21.5-30 Hz) and Gamma (30.5-44 Hz) band power over the right temporal cortex [47].

Taken together, these studies highlight the potential of single rTMS sessions in provoking neuroplasticity or rather in modulating tinnitus-related ongoing brain activity [26,31,32]. However, some methodological shortcomings should be noted. None of them performed rTMS with the aid of a neuronavigation system, which enables individual cortical targeting together with precise and inter-session reliable coil placement. Cap-based coil placement was identified as an inherent factor for target variability [48]. Moreover, the available studies did not incorporate a sham stimulation which adequately mimics the sensory sensations of a verum stimulation. Furthermore, high frequency stimulations such as 10 Hz or 20 Hz have not been systematically investigated in previous studies, even if such protocols applied to the left or right temporal cortex have also been reported to evoke short-term tinnitus suppressions [42,44,45,49–51].

With the present experiment we wanted to contribute to the rTMS-EEG research branch and overcome these identified shortcomings. Hence, we aimed for an investigation of the effects of low and high frequency rTMS (1 Hz, 10 Hz, 20 Hz) compared to an active sham condition of 0.1 Hz (no neuroplastic effects [52,53]) applied with an e-field guided neuronavigation system to the left and right temporo-parietal junction (TPJ) on tinnitus loudness and oscillatory brain activity. The TPJ was identified as the most effective target for rTMS in tinnitus [35]. Further we opted for the identification of a patient-specific rTMS protocol for maximum and sham-superior transient tinnitus loudness suppression akin to previous studies [44] and likewise evaluated its consequences on resting state EEG activity. Particularly, a focus on protocols which induce strongest loudness reduction has the potential to uncover potential oscillatory biomarkers for tinnitus suppression.

## II. Materials & methods

The dataset of tinnitus patients reported in the present study as well as all experimental proceedings have already been described in Schoisswohl et al., 2021 [43], and are cultivated for the purpose of analyzing potential rTMS-induced changes in spontaneous brain activity.

### A. Participants

22 patients (five female) with chronic subjective tinnitus (> six months) were recruited at the Interdisciplinary Tinnitus Center in Regensburg, Germany. Inclusion criteria for the present experiment were: age between 18 and 75 years; German-speaking; no severe somatic, neurological or psychiatric condition such as depression, acute psychosis, substance abuse, known brain tumor or encephalitis plus no or stable medication with psychoactive drugs. In the case of any contraindication with respect to TMS (e.g., epilepsy or condition after traumatic brain injuries) as well as magnetic resonance imaging (MRI) (e.g., heart pacemaker, other metallic/electrical body implants such as insulin pumps, or severe claustrophobia), study participation was not permitted. Further, a parallel participation in other tinnitus-related studies or treatments was not allowed.

### B. Experimental procedure

All experimental proceedings were reviewed and approved by the local ethics committee of the University of Regensburg, Germany (ethical approval number: 17-820-101). Each patient was carefully informed about the study’s purpose, methodological approaches and potential adverse effects of TMS. Prior participation, each patient provided written informed consent.

All patients underwent clinical audiometry (125 Hz to 8 kHz), performed with a Madsen Midimate 622D audiometer (GN Otometrics, Taustrus, Denmark). Anatomical T1-weighted MRI scans were obtained with a MAGNETOM 1.5 Tesla scanner (Siemens, Munich, Germany) for neuronavigated TMS. Further, patients had to fill out the Tinnitus Handicap Inventory [54,55], the Tinnitus Sample Case History Questionnaire (TSCHQ; [56]) and the European School for Interdisciplinary Tinnitus Research Screening Questionnaire (ESIT-SQ; [57]).

The actual experiment consisted of the application of eight different short-term magnetic stimulations sessions, whereby the same four rTMS protocols were applied over the left and right TPJ (see section *Repetitive Transcranial Magnetic Stimulation*). The hemisphere to start with as well as the order of applied stimulation protocols was randomized per patient. Before and after each stimulation protocol, patient’s spontaneous brain activity was recorded for three minutes by EEG (see section *Electroencephalography - recording and preprocessing*). For this, patients were instructed to sit still and avoid unnecessary muscle movements and eye blinks/movements as good as possible plus focus on a particular point in the room. In parallel to pre and post rTMS EEG recordings, patients were requested to evaluate the current loudness of their tinnitus perception on a visual analogue scale anchored from 0% (complete absence of tinnitus) over 100% corresponding to the patient’s typically perceived tinnitus loudness to 110% (10% increase compared to the typically perceived tinnitus loudness) in 10% steps. The rating was conducted over a period of 3 minutes at seven points in time every 30 seconds before (T0_pre_-T180_pre_) and after each stimulation protocol (T0_post_-T180_post_). Between each protocol a minimum interval of six minutes was kept (three minutes of EEG recording before and after rTMS) – if necessary, the interval was extended until the patient reported a return to the originally perceived tinnitus loudness.

### C. Repetitive transcranial magnetic stimulation (rTMS)

All magnetic stimulations were performed with an e-field guided TMS system (NBT System 2; Nexstim Plc., Helsinki, Finland) with an orientation of the induced e-field perpendicular to the sulcus of the stimulated brain area. This was enabled by a co-registration of individual anatomical brain scans (T1-weighted) and an online visualization of strength (V/m) and orientation of the induced e-field on a patient-specific 3D head model.

TMS stimulation intensity was based on patient’s resting motor threshold (RMT) and determined before experiment start. Thereby, various positions of the left primary motor cortex were stimulated until several motor evoked potentials (MEPs) with a peak-to-peak amplitude greater than 50 µV, recorded from the thenar muscles of the right hand via system built-in Electromyography, became apparent. The stimulation position, which exhibited the highest response (motor hotspot), was repeated via a system-intern aiming tool (same coil orientation and tilting) and the RMT was defined by the system-integrated maximum likelihood threshold hunting algorithm with mounted EEG cap. The RMT was defined as the lowest stimulation intensity required to elicit MEPs with a peak-to-peak amplitude of at least 50 µV in at least 50% of the applied pulses.

Over the course of the experiment, three different active and one sham rTMS protocol were administered over the left and right hemisphere in a randomized order. For active magnetic stimulation 200 pulses of 1 Hz, 10 Hz and 20 Hz rTMS were applied with an intensity of 110 % RMT over the left and right TPJ. The electrode positions CP5 and CP6 (10-20 system) served as reference points for TPJ localization. For sham stimulation, an active control condition was used which consisted of 20 pulses of 0.1 Hz rTMS at an intensity of 110% RMT administered over the left and right TPJ as well. 0.1 Hz rTMS is assumed to not evoke neuroplasticity [52,53]. Previous rTMS-EEG studies [46,47] lacked of implementing a sham stimulation (45° coil angulation; backside of coil over CPz) which closely reproduces the sensory sensation of verum rTMS. Hence, a total number of eight different magnetic stimulation protocols were applied per patient (4 protocols × 2 hemispheres). Since (temporary and often minor) rTMS-changes in tinnitus loudness are relatively difficult to evaluate, we used an uncooled figure-of-eight coil for all stimulations to refrain from producing additional noise by the coil cooling system. As TMS is typically accompanied by loud click noises, each patient had to wear in-ear plugs to prevent hearing damage. Detailed information about the used procedures can be found in previous work [43,45].

### D. Electroencephalography – recording and preprocessing

Before and after each magnetic stimulation, resting state EEG was recorded for three minutes with a BrainAmp DC system and an EasyCap electrode cap with 64 electrodes placed according to the 10-20 system (GND: AFz, REF: FCz) together with the software Brain Vision Recorder 1.20 (Brain Products GmbH, Germany). Sampling rate was 500 Hz. Impedances were kept below 10 kΩ for the entire recording.

The recorded EEG data was down-sampled to 250 Hz after being low-pass filtered at 500 Hz to avoid aliasing during the down-sampling process. Next, the signals were filtered using a linear-phase Finite Impulse Response (FIR) filter with cutoff frequencies of 0.1 Hz and 100 Hz to retain relevant frequency content across various bands. A zero-phase FIR notch filter with a center frequency at 50 Hz and stop/transition bands of 0.25 Hz and 1 Hz, respectively, was applied to eliminate electrical interference or line noise at 50 Hz. Each channel was then re-referenced by averaging the signal from the channel itself. Independent Component Analysis (ICA) was used to separate the EEG data into components, and eye movement and blink-related components were automatically identified and removed using the *MNE-ICALabel* Python package [58], provided the prediction score exceeded 70% and the component was labeled as horizontal or vertical eye movement.

### E. Behavioral data analysis

Behavioral analyses were carried out using the statistic software R (R version 4.2.3; R Foundation for Statistical Computing, Austria) together with the packages “psych”, “emmeans, “sjstats”, “lme4” and “ggplot2”. Tinnitus loudness ratings before each magnetic stimulation protocol (T0_pre_-T180_pre_) were averaged to receive a stable baseline loudness level for individual subjects per protocol. This baseline was then cultivated to calculate TMS-induced loudness changes (Δloudness = post – pre) for each stimulation protocol and rating timepoint after stimulation (T0_post_-T180_post_). To evaluate potential differences between timepoints as well as among stimulation protocols, a linear mixed effect model was used with the dependent parameter Δloudness and the fixed effects *Time* (T0_Δ_, T30_Δ_, T60_Δ_, T90_Δ,_ T120_Δ_, T150_Δ_, T180_Δ_), *Protocol* (left and right 1, 10, 20, 0.1 Hz) and the interaction *Time* × *Protocol*. The individual subject was considered as random effect in the model. Fixed effects were evaluated with the Expected Mean Square Approach. In case of significant results, post hoc Tukey-tests were performed to assess differences within fixed effects. The level of significance was set to p ≤.05 for all analyses.

### F. Electroencephalography data analysis

All electrophysiological data were analyzed using Python (vers. 3.10.9) with the MNE package (vers. 1.8) [59].

#### 1. Power analyses and source reconstruction

Average power for each individual channel was computed across five distinct frequency bands: Delta (0.5–4 Hz), Theta (4–8 Hz), Alpha (8–13 Hz), Beta (13–30 Hz), and Gamma (30–80 Hz). This calculation was carried out using the Welch method for spectral estimation [60] in which the signal was divided into overlapping segments, Hanning window function was applied to each segment to minimize edge effects, and the periodogram for each segment was computed. These periodograms were then averaged to estimate the power spectral density (μV^2^/Hz). To estimate the forward operator for our EEG recordings, we employed a boundary element model, source model, and co-registrated data from a standard template MRI subject provided by the *MNE* Python package [59]. Since no specific data period was available to estimate noise covariance in the EEG recordings, we used an ad hoc covariance matrix to model sensor noise. This matrix approximates Gaussian noise on the sensors, assuming an infinite number of samples, making it a practical solution when direct noise estimation is not possible. Using this noise covariance matrix along with the forward model, we applied the linear minimum-norm inverse method dynamic Statistical Parametric Mapping (dSPM) to calculate the inverse solution [59]. This provided us with source time courses and power spectra. For each brain label, we obtained a single time course by averaging across vertices at each time point within the label. The brain labels and cortical parcellation were derived from the Desikan-Killiany Atlas [61].

#### 2. Statistical analyses of spectral power

Pre- to post-stimulation changes in EEG power spectra were analyzed using dependent-samples t-tests for each verum stimulation protocol, across frequency bands and brain vertices/labels. Statistically significant changes were further contrasted against the spectral power change (Δpower = post - pre) induced by the sham protocol applied to the same hemisphere as the respective verum stimulation protocol (e.g., left 1 Hz vs. left 0.1 Hz; right 10 Hz vs. right 0.1 Hz). We were solely interested in brain regions which demonstrated significant and sham-superior power spectra changes from pre to post stimulation. Pearson correlations were used to evaluate potential linkages with behavioral changes in tinnitus loudness (Δloudness = post - pre) and brain regions showing significant and sham-superior power spectra changes.

We were further interested in assessing differences between rTMS protocols with respect to oscillatory brain activity changes. Therefore, we defined a region of interest (ROI) around the TPJ for the left and right hemisphere. The ROI which was comprised of the superior, middle and transverse temporal gyri, the inferior and superior parietal gyri and the supramarginal gyrus. We averaged power spectra changes for this ROI per protocol (left hemisphere ROI - left hemispheric protocols; right hemisphere ROI - left hemispheric protocols) and contrasted them with dependent sample t-tests. The level of significance was set to p ≤.05 for all EEG analyses as well.

#### 3. Connectivity

To compute spectral connectivity between brain regions, we segmented the data into 2-second epochs and used a continuous wavelet transform with Morlet wavelets (7 cycles, zero mean) for the analysis. Connectivity was assessed using two different methods: the weighted phase lag index (wPLI) [62], which estimates whether the signal in one brain region leads or lags behind the signal in another region, and coherence, which measures the absolute value of the complex correlation between signals in the frequency domain [63]. Dependent t-tests were conducted for each edge (connection between two brain regions) to compare pre to post spectral connectivity changes. Given the high number of comparisons, we applied multiple-testing corrections to the connectivity results using both the Bonferroni correction and the Benjamini–Hochberg false discovery rate procedure. Connectivity analysis was done using the MNE-connectivity package (vers. 0.7.0) [59] in Python.

### G. dentification of the individual best protocol, corresponding EEG spectral power changes & correlation with tinnitus suppression

Based on the premise of intra- and inter-subject variability in responses to rTMS protocols [43–45], we were interested in identifying a protocol evoking strongest and sham-superior suppression of the tinnitus percept in individual subjects. For this purpose, we evaluated average tinnitus loudness ratings (T0-T180) before and after each magnetic stimulation protocol and in case of a suppression (Δloudness = post - pre) we checked if this was superior to the sham stimulation of the respective hemisphere (e.g., Δloudness right 10 Hz > Δloudness right 0.1 Hz) in the individual subject. We used the behavioral data of the individual subject’s best protocol for additional analysis of pre to post power changes as well as to correlate EEG and tinnitus loudness changes. Though, exclusively for the Alpha and Gamma power spectra, as these frequency bands seem to be most relevant for tinnitus loudness changes [29,32,46,64–68].

## III. Results

### A. Sample characteristics

Tinnitus patients were between 43 and 69 years old (M = 57.04 ± 6.79), reported to perceive tinnitus for 131.65 months on average (SD = 116.76) and yielded a mean THI sum score of 58.49 (SD = 19.55), which represents severe tinnitus handicap (grade 4). The mean RMT (%) and induced e-field (V/m) with mounted EEG cap were 41.91 (SD = 9.65) and 66.66 (SD = 15.38), respectively. Mean hearing loss was 23.42 dB (SD = 9.72) for the left and 27.87 dB (SD = 14.29) for the right ear.

### B. Behavioral results

No effect of *Time* or an interaction of *Time* × *Protocol* was evident. Protocol-specific changes in tinnitus loudness are illustrated in **Figure 1 A** and **B** for all evaluation timepoints (T0-T180) and protocols. We observed a significant effect of *Protocol*. Ensuing post hoc contrasts revealed significant differences between right 10 Hz versus right 1 Hz, left 10 Hz, left 0.1 Hz, right 0.1Hz; right 20 Hz versus left 10 Hz, left 0.1 Hz, right 0.1Hz; left 20 Hz versus 0.1 Hz left and left 1 Hz versus left 0.1 Hz. Thereby, the first mentioned protocol always demonstrated higher tinnitus loudness suppression. On group level, right 10 Hz induced the strongest tinnitus loudness suppression. An overview of induced tinnitus loudness suppression per protocol plus comparisons between protocols can be found in **Figure 1 C**.

**Figure 1.**
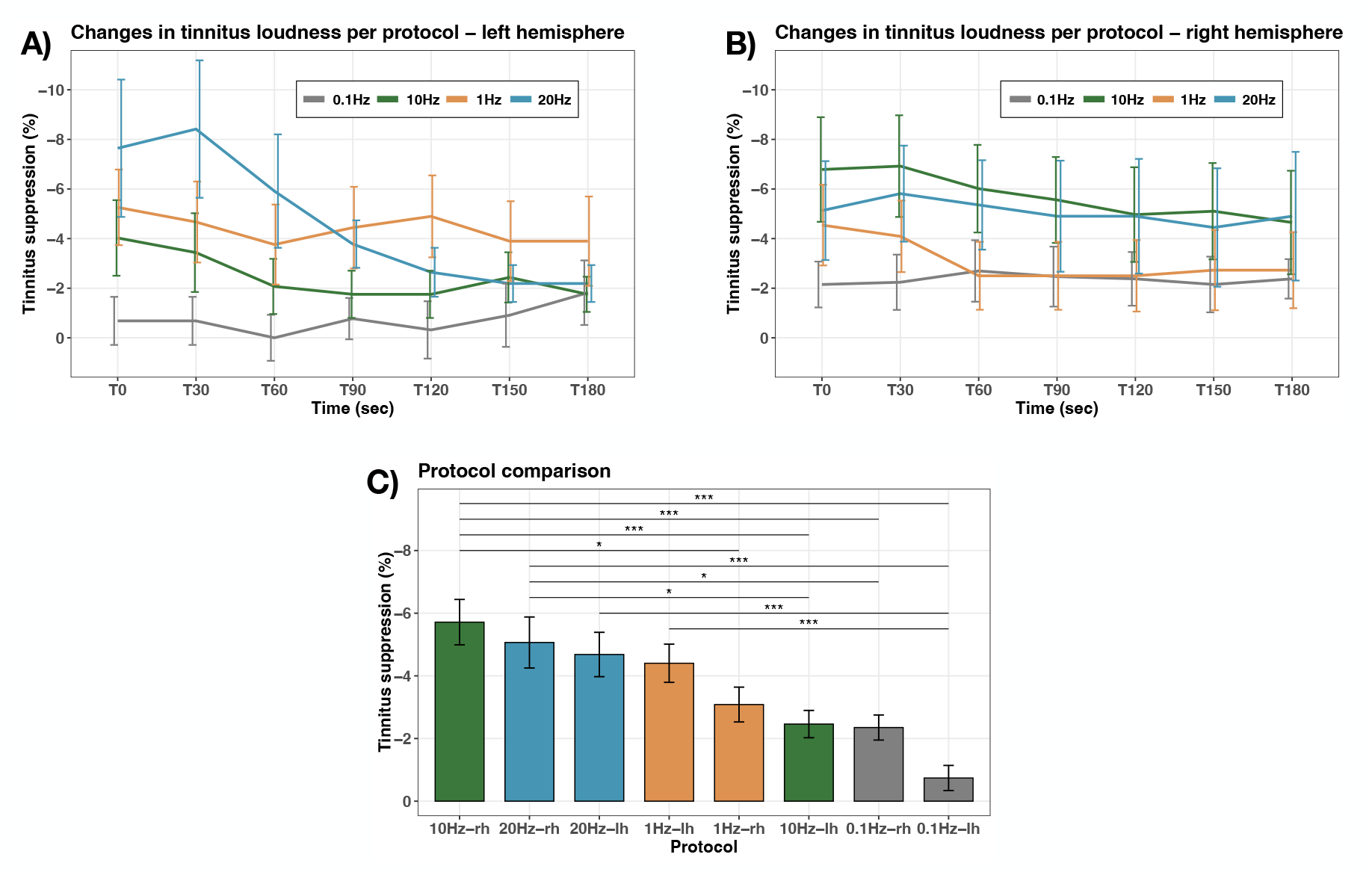
Tinnitus suppression per protocol & protocol comparisons. Tinnitus loudness changes from pre to post magnetic stimulation are illustrated for each protocol for the **A)** left hemisphere and for the **B)** right hemisphere at all seven rating points (T0-T180). **C)** Differences between protocols with respect to induced tinnitus loudness suppression. Significant differences are highlighted with asterisks. Error bars indicate standard errors. lh = left hemisphere; rh = right hemisphere; * p ≤.050; ** p ≤.010, *** p ≤.001.

### C. Electroencephalography results

At the vertex level, no significant effects were detected using the cluster-based permutation approach. Significant and sham-superior power spectra changes were observed for the protocols right 10 Hz, left 20 Hz and right 20 Hz. A detailed overview of brain regions demonstrating sham superior power spectra changes can be found in **Figures 2** and **3** separated per rTMS protocol and frequency band. 10 Hz stimulation over the right TPJ resulted in increased activity in the Alpha frequency range in the occipital cortex and medial temporal lobe of the left hemisphere (**2A**). Further increased Gamma activity was observed in parts of the parietal lobe and the anterior section of the temporal cortex of the right hemisphere (**2B**). A 20 Hz stimulation of the left TPJ resulted in an increase of Gamma power exclusively in the right fusiform gyrus (**2C**).

**Figure 2.**
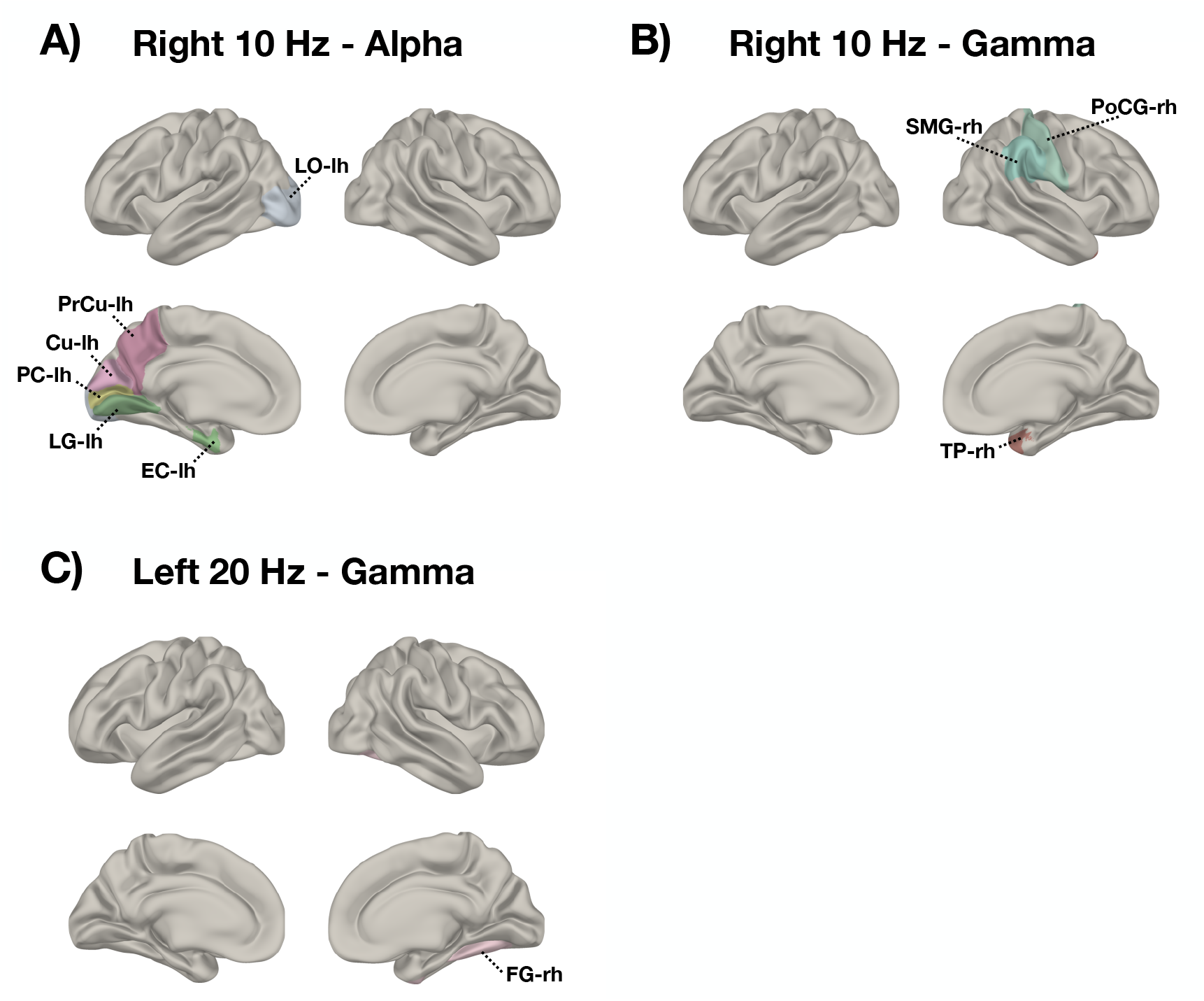
Significant and sham-superior changes in EEG power spectra (µV2/Hz) – right 10 Hz & left 20 Hz. Brain labels demonstrating statistically significant and sham-superior power spectra increases in the A) Alpha (8-13 Hz) and B) Gamma (30-80 Hz) frequency bands following 10 Hz rTMS of the right TPJ and in the C) Gamma (30-80 Hz) band after 20 Hz rTMS over the left TPJ. lh = left hemisphere; rh = right hemisphere; LO = lateral occipital cortex; PrCu= precuneus; Cu = cuneus; PC= pericalcarine cortex; LG = lingual gyrus; EC = entorhinal cortex; SMG = supramarginal gyrus; PoCG = postcentral gyrus; TP = temporal pole; FG = fusiform gyrus.

**Figure 3.**
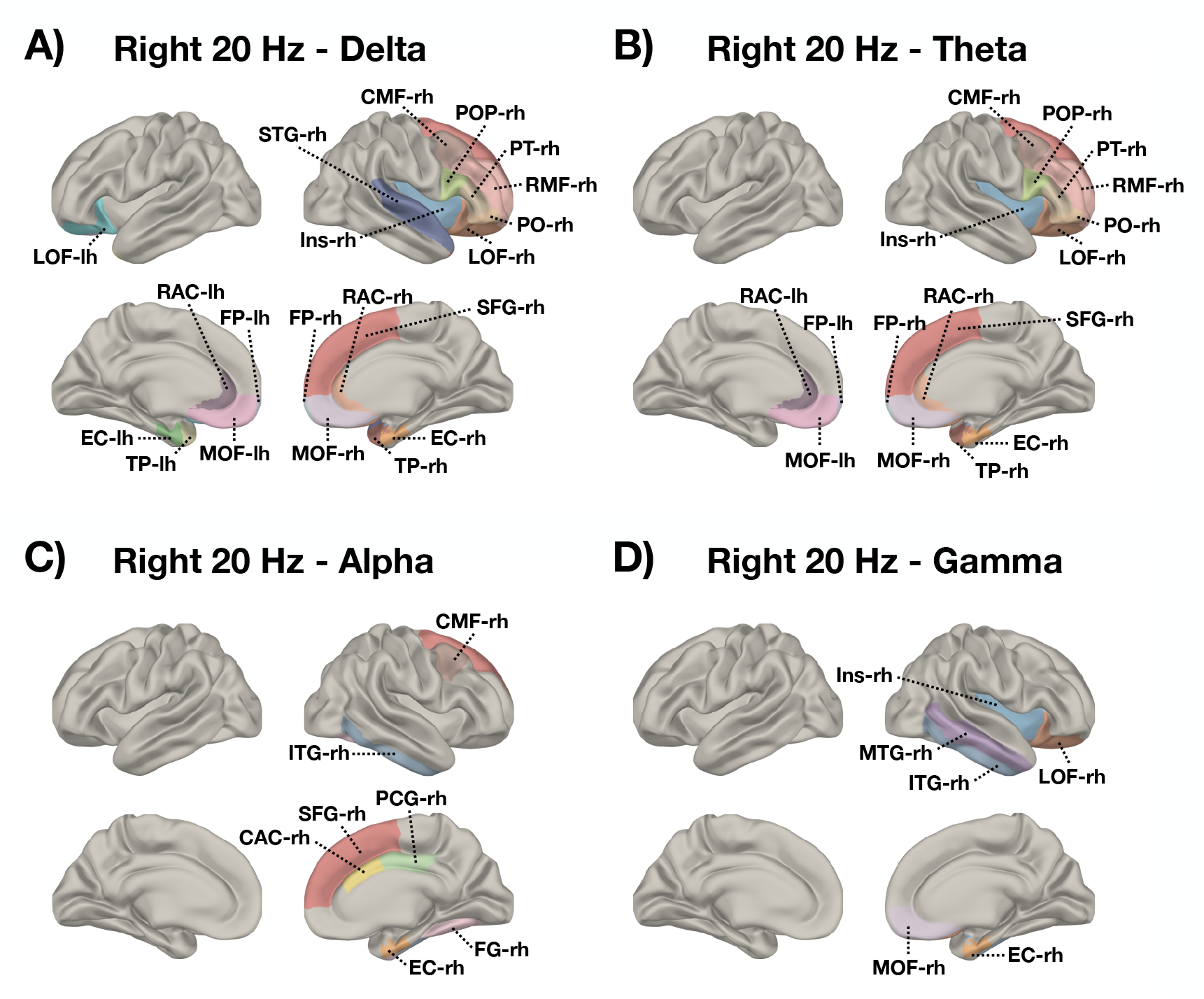
Significant and sham-superior changes in EEG power spectra (µV^2^/Hz) – right 20 Hz. Brain labels demonstrating statistically significant and sham-superior power spectra increases in the **A)** Delta (0.5-4 Hz) and **B)** Theta (4-8 Hz) **C)** Alpha (8-13 Hz) and **D)** Gamma (30-80 Hz) frequency band following a stimulation of the right TPJ with 20 Hz rTMS. lh = left hemisphere; rh = right hemisphere; LOF = lateral orbitofrontal cortex; RAC = rostral anterior cingulate cortex; FP = frontal pole; MOF = medial orbitofrontal cortex; TP = temporal pole; EC = entorhinal cortex; STG = superior temporal gyrus; CMF = caudal middle frontal gyrus; POP = pars opercularis; PT = pars triangularis; RMF =rostal middle frontal gyrus; PO = pars orbitalis; Ins = Insula; SFG = superior frontal gyrus; ITG = inferior temporal gyrus; CAC = caudal anterior cingulate cortex; PCG = posterior cingulate cortex; FG = fusiform gyrus; MTG = middle temporal gyrus.

Changes in Delta, Theta, Alpha and Gamma power spectra were detected after right hemispheric 20 Hz rTMS. Increases in Delta frequency band power appeared in frontal as well as temporal regions of both hemispheres (**3A**). Enhanced Theta power was available in frontal brain areas of both hemispheres as well as in right medial temporal parts (**3B**). In right frontal and temporal cortices, we identified elevated power in the Alpha range (**3C**). Gamma band activity increased from pre to post stimulation in parts of the right frontal cortex and the right temporal gyrus (**3D**). Power values before and after stimulation can be seen from the Supplementary Material for each identified protocol per frequency band and brain label (**Figures S1** and **S2**).

No significant connectivity changes from pre to post stimulation was present in any protocol. No correlations between changes in behavioral and electrophysiological metrics were available.

Comparisons of induced EEG power spectra changes among all protocols using a predefined ROI revealed significant differences between left TPJ sham rTMS (0.1 Hz) and 10 Hz (t = −2.38, p =.027) and 20 Hz (t = 2.16, p =.043) in the Delta respectively Alpha frequency bands in favor of the verum protocols.

### D. Identification of the individual best protocol, corresponding EEG spectral power changes & correlation with tinnitus suppression

In 16 subjects it was feasible to identify a personalized protocol which elicited sham-superior tinnitus loudness reduction. For the remaining 6 subjects, none of the applied protocols evoked a sham-superior suppression. For four subjects each, stimulation at 10 Hz over the right TPJ and 20 Hz over the left TPJ produced the strongest suppression. In three participants each, 1 Hz over either the left or right hemisphere showed the strongest change in tinnitus loudness. One subject each, reported left 10 Hz or right 20 Hz to induce the most pronounced suppression. These protocols resulted in a mean loudness suppression of 16.88% (SD = 9.40). For a detailed overview of the average tinnitus loudness suppression per protocol and subject see **Table S1** in the Supplementary Material.

A stimulation with the individual best protocol resulted in a significant increase in Alpha power in frontal brain areas (see **Figure 4A**). Pre and post stimulation EEG power values for best protocols can be found in **Figure S3** in the Supplementary Material. We observed significant positive correlations between changes in tinnitus loudness and changes in EEG power spectra exclusively in the Alpha frequency range (*r*_*min*_ =.51; *r*_*max*_ =.65; *p*_*min*_ =.007; *p*_*max*_ =.043). Accordingly, an increase in tinnitus suppression is related to an increase in Alpha activity. EEG and behavioral correlations were predominately observed in frontal areas. Brain regions exhibiting correlations are highlighted in **Figure 4B**.

**Figure 4.**
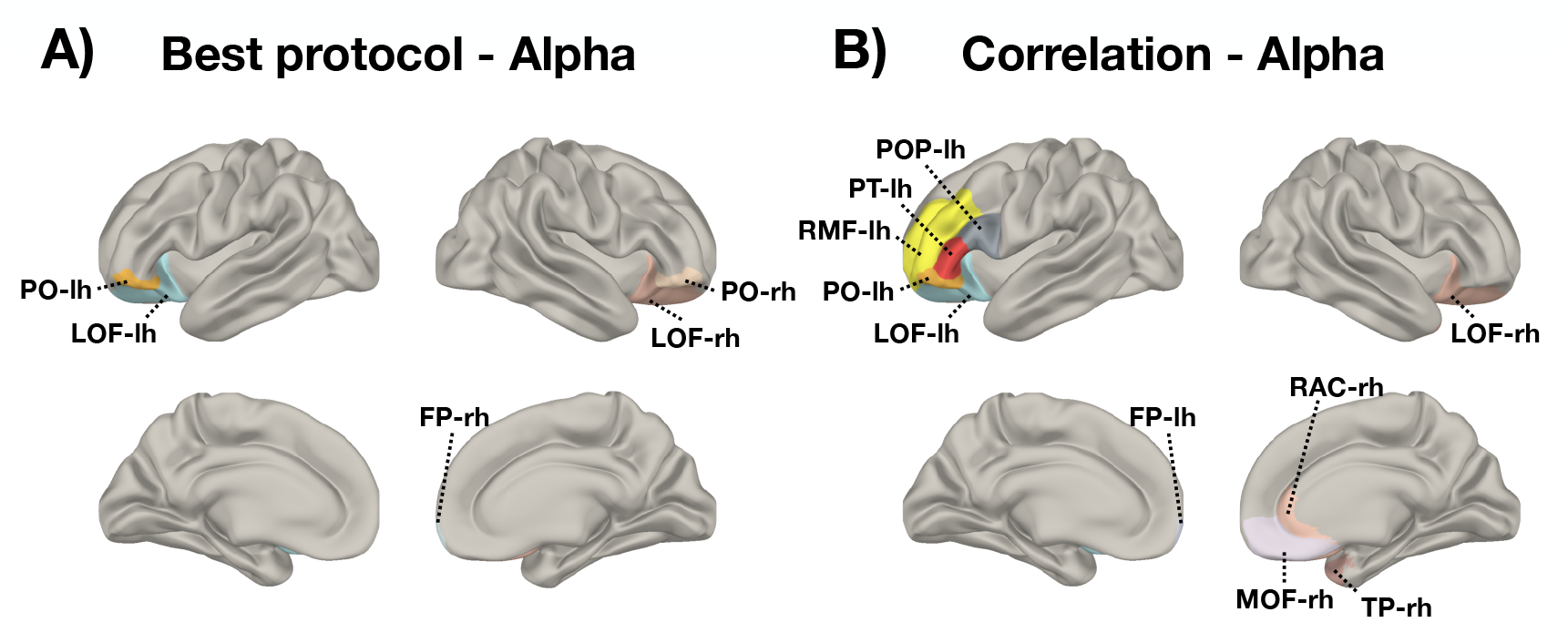
Significant changes in EEG power spectra (µV^2^/Hz) for individual best protocol & correlation with tinnitus suppression: A) Brain labels with significant increases from pre to post stimulation following a stimulation with the individual best protocol for sham-superior and maximum tinnitus loudness suppression and **B)** significant correlations (r_min_ =.51; r_max_ =.65; p_min_ =.007; p_max_ =.043) with transient suppression of the tinnitus percept are illustrated for the Alpha (8-13 Hz) frequency band. lh = left hemisphere; rh = right hemisphere; FP = frontal pole, LOF = lateral orbitofrontal cortex, PO = pars orbitalis, PT = pars triangularis, RMF =rostal middle frontal gyrus, TP = temporal pole, RAC = rostal anterior cingulate cortex, MOF = medial orbitofrontal cortex, POP = pars opercularis.

## IV. Discussion

The present experiment sought to investigate potential modulations in resting state EEG activity and changes in tinnitus loudness following various brief rTMS protocols (0.1 Hz, 1 Hz, 10 Hz, 20 Hz) applied over the left and right temporo-parietal junction (TPJ) using an e-field guided neuronavigation system. Furthermore, we examined the effects of rTMS protocols that elicited the strongest and sham-superior transient tinnitus loudness suppression in the individual patient on EEG activity, as this may help to uncover potential oscillatory biomarkers of tinnitus suppression.

### A. Behavioral results

Akin to previous studies, rTMS protocols with a modest amount of applied pulses are capable to induce short-term suppressions of the tinnitus percept [41–45,49]. Traditionally left-hemispheric 1 Hz rTMS is applied in tinnitus treatment [1,37,40]. In our study this protocol ranked only fourth in efficacy, whereas high frequency stimulations, in particular 10 Hz of the right TPJ, evoked strongest effects. While high frequency protocols are generally thought to increase cortical excitability [14], tinnitus-related pathological brain activity might alter or reverse stimulation effects and potentially shifts them towards inhibition. Such state-dependent mechanisms could potentially explain our observation of high frequency rTMS superiority in alleviating patient’s tinnitus loudness. Moreover, this finding somehow implicates that alternative protocols could potentially yield greater therapeutic benefits - high frequency rTMS over temporal regions is already evaluated in treatment schedules with promising results [69,70]. Except for right-hemispheric 1 Hz and left-hemispheric 10 Hz, each protocol evoked suppressions superior to sham stimulation (0.1 Hz) of the corresponding hemisphere. Given these findings, TMS can be considered an effective approach for brief tinnitus suppression as it consistently outperforms placebo stimulation.

### B. Electroencephalography results

#### 1. Effects of the different investigated protocols

According to several neurophysiological studies, tinnitus is characterized by enhanced oscillatory activity in Delta, Theta and Gamma frequency bands and reduced activity in the Alpha frequency range. For example, Alpha is assumed to be pathologically reduced in tinnitus patients due to deficient inhibitory mechanisms and the persistent perception of a phantom sound, leading to enhanced Gamma activity, which has also been discussed as an oscillatory signature of tinnitus [24–28,64,65,71]. Insights from acoustic stimulation experiments suggest that during the state of brief tinnitus suppression a temporary inversion of pathological brain activity takes place [66,72]. Thus, rTMS protocols that attenuate the tinnitus percept are expected to modulate tinnitus-related brain activity patterns as well e.g., by reducing Gamma and/or increasing Alpha power.

Interestingly, we observed significant and sham-superior increases in Delta (0.5–4 Hz), Theta (4–8 Hz), Alpha (8–13 Hz) and Gamma (30–80 Hz) power spectra exclusively in the top three protocols inducing sham-superior suppression of the tinnitus percept (right 10 Hz, right 20 Hz, left 20 Hz). Increases in Gamma band power were most pronounced in frontal, temporal as well parietal cortical regions (right 10 Hz, left 20 Hz, right 20 Hz). This observation is not only reflected in the results by Müller et al., who report an decrease frontal Gamma linked to an elevation of tinnitus loudness after rTMS [46], but also supported by insights from acoustic stimulation experiments which likewise observed an increase in Gamma activity during brief acoustic tinnitus suppression [67,73,74]. Hence, these observations challenge the notion that pathologically increased Gamma activity is related to tinnitus perception.

Delta and Theta power increases were strongest in frontal and temporal regions (right 20 Hz). These frequency bands have been reported to be elevated in tinnitus compared to healthy controls [25,75,76] and to be reduced during brief acoustic tinnitus suppression [72,73]. Our observation is also in contrast to the findings of a previous rTMS-EEG study in tinnitus by Schecklmann et al., who could show decreases in Delta and Theta band power in frontal sensors following left temporal 1 Hz [47]. However, in the present analyses left temporal 1 Hz did not evoke any significant and sham-superior modulations in EEG activity.

In accordance with the prevalent electrophysiological models for tinnitus, we found an Alpha power increase in frontal, temporal and occipital regions (right 10 Hz, right 20 Hz). Equally, Müller et al. reported that rTMS-induced tinnitus suppression is connected to an Alpha power increase in the auditory cortex [46]. In contrast to healthy controls, activity in the Alpha range is decreased in tinnitus patients, potentially due to deafferentation and diminished inhibitory processes [24,25,32,71,77]. This finding of an inversion of pathological Alpha activity in temporal and frontal regions may reflect a temporary restoration of inhibitory and excitatory dynamics during tinnitus suppression and the reduced processing of patient’s tinnitus percept. Both regions play a key role in the perception of auditory sensations, including tinnitus [19,78,79].

Taken together, the present results highlight the potential of rTMS, particularly high frequency protocols, to modulate tinnitus-related oscillatory brain activity. In line with basic science evidence [5,7–10,80] our results emphasize the capacity of rTMS to evoke widespread changes in cortical excitability and neuroplasticity, potentially reflecting network activity beyond the auditory cortex that contributes to tinnitus suppression. Pathological changes in tinnitus have also been reported in brain regions outside the auditory system [79,81– 83]. It should be noted that we did not observe any connectivity changes following rTMS. Even though the observed neuroplastic effects accompanied the strongest and sham-superior tinnitus reduction (on group-level), these should be interpreted with caution, as EEG and behavioral changes were uncorrelated, akin to previous studies [46,47,66,72]. However, our data highlights the importance of rTMS targets beyond the auditory cortex. Stimulations of non-auditory regions are already successfully applied in single sessions aiming for brief suppressions [44,78] as well as treatment schedules e.g., high-frequency rTMS of the frontal cortex to target the affective component associated with tinnitus [78,84,85].

#### 2. Individual best protocol

Based on the premise of intra- and inter-subject effect variability, we aimed to identify a protocol evoking maximal and sham-superior tinnitus suppression for each tinnitus patient with the aim to examine its consequences on resting state EEG activity. Such a personalized protocol might provide insights behind the neurophysiological mechanisms of effective tinnitus suppression and may help to uncover oscillatory biomarkers associated with tinnitus loudness modulation. This was achievable in 73% of tinnitus patients. Personalization of rTMS protocols provide a promising avenue to enhance therapeutic effects [44,86].

A stimulation with the individual best protocol caused an increase in power spectra exclusively for the Alpha frequency band in left and right frontal areas. Changes in tinnitus loudness were positively correlated with an increase in frontal Alpha power (↓ tinnitus loudness; ↑ Alpha power). In a resting state functional MRI study it was demonstrated that activity in frontal areas is negatively correlated with tinnitus loudness [87]. Vanneste et al. [88] analyzed ongoing brain activity in a unique sample of tinnitus patients with intermittent tinnitus characterized by certain days with and other days without tinnitus perception. A contrast between these conditions also revealed increased Alpha activity in frontal regions during tinnitus off-states. The frontal cortex is involved in the integration of multi-sensory information as well as the emotional processing of sounds and plays a decisive role in the top-down modulation of automatic or peripheral physiological reactions to emotional events [79,87]. In this context, the observed increase in frontal Alpha power might be indicative of enhanced inhibitory processes in the frontal cortex potentially reflecting reduced emotional and salient processing of the tinnitus percept during brief tinnitus suppression. Thus, frontal Alpha activity may serve as an oscillatory marker for tinnitus perception respectively suppression.

Altogether our findings of a correlation between reported loudness reduction and Alpha power increase for the individually best protocol supports an individualized treatment approach: in different patients, different protocols evoke the best transient tinnitus suppression and only with these protocols we find a correlating change in neuronal oscillatory activity.

### C. Limitations

In the interpretation of the present findings some shortcomings should be considered. Changes in tinnitus loudness were relatively small and could have been difficult for the patients to rate. With 200 applied pulses, stimulation protocols were relatively short. A larger number of administered pulses could potentially induce stronger and unambiguous short-term suppressions of patient’s tinnitus percept. More sophisticated sham conditions beyond 0.1 Hz mimicking the sensory sensation of TMS by e.g., electrical stimulation of the scalp [89,90], may provide a better control condition and yield more robust findings. Even though patients wore ear protection and we used an uncooled coil, the loud click noise of TMS pulses could have biased our findings. No fully silent TMS system is available. TMS coils with reduced acoustic noise are already under development [91]. We used a neuronavigation system to ensure precise cortical targeting. Though, head movements throughout stimulation may have reduced target precision. Future studies could benefit from robot-assisted neuronavigated TMS, which automatically compensates for head movements [80,92]. Further, our findings were not corrected for multiple comparisons and should therefore be interpreted as exploratory.

## V. Conclusion

The aim of the present study was to investigate potential changes in resting state EEG and tinnitus loudness following several brief rTMS protocols applied over the temporo-parietal junction. With the present findings we can demonstrate that single session rTMS effectively induces short-term suppressions of the tinnitus percept and evokes modulations of tinnitus-related oscillatory brain activity dynamics. In examining the individual optimal protocol for tinnitus suppression, our findings indicate that increased activity in the Alpha frequency range localized in the frontal cortex may serve as a potential oscillatory marker for effective tinnitus suppression.

## Data Availability

All data produced in the present study are available upon reasonable request to the authors

## Ethics approval and consent to participate

The study was conducted according to the guidelines of the Declaration of Helsinki and approved by the Ethics Committee of the University of Regensburg, Germany (ethical approval number: 17-820-101). Informed consent was obtained from all subjects involved in the study.

## AVAILABILITY OF DATA AND MATERIALS

The datasets used and/or analyzed during the present study are available from the corresponding author on reasonable request.

## CODE AVAILABILITY

All code used for EEG processing and creating the figures are openly available at: https://github.com/payamsash/regTMS.

## Acknowledgment

We would like to thank Susanne Staudinger, Viola Lents and Leonie Forstner for their valuable help in experiment management and data acquisition. We thank all participants of the study for their time and effort. Further, we want to thank Nexstim Plc. for providing the electric-field guided neuronavigation rTMS system to perform this experiment.

## DECLARATION OF AI USE

ChatGPT-4o has been used for AI-assisted copy editing.

## AUTHOR’S CONTRIBUTION

**Stefan Schoisswohl:** Conceptualization, Data Curation, Formal Analyses, Investigation, Methodology, Project Administration, Writing - Original Draft; **Patrick Neff:** Supervision, Writing - Review & Editing; **Martin Schecklmann:** Conceptualization, Funding Acquisition, Methodology, Project Administration, Resources, Supervision, Writing - Review & Editing; **Berthold Lannguth:** Funding Acquisition, Resources, Supervision, Writing - Review & Editing; **Tobias Kleinjung:** Writing - Review & Editing; **Payam S. Shabestari:** Data Curation, Formal Analyses, Methodology, Writing - Original Draft

---

**Figure S1.**
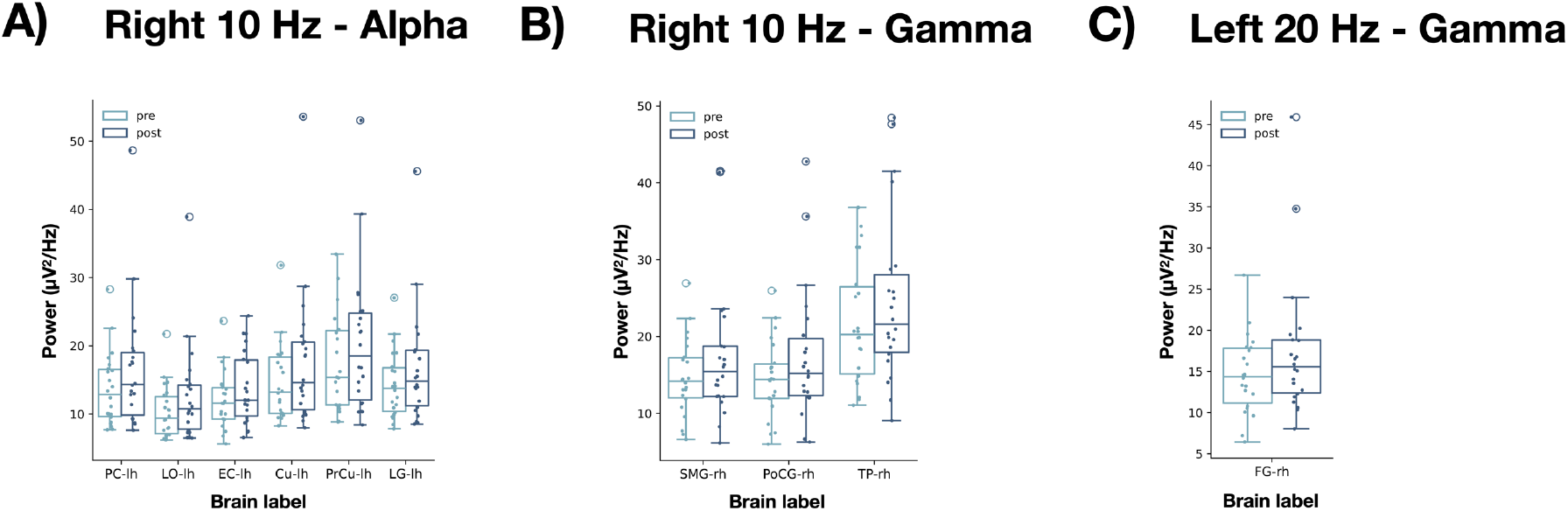
Pre and post EEG power values (µV^2^/Hz) for right 10 Hz and left 20 Hz rTMS Figure S1. Boxplots illustrate pre and post stimulation EEG power values (µV^2^/Hz) for identified brain labels with a statistically significant and sham-superior power spectra increase in the **A)** Alpha (8-13 Hz) and **B)** Gamma (30-80 Hz) frequency bands following 10 Hz rTMS of the right TPJ and in the **C)** Gamma (30-80 Hz) frequency band after a stimulation with 20 Hz over the left TPJ. lh = left hemisphere; rh = right hemisphere; LO = lateral occipital cortex; PrCu= precuneus; Cu = cuneus; PC= pericalcarine cortex; LG = lingual gyrus; EC = entorhinal cortex; SMG = supramarginal gyrus; PoCG = postcentral gyrus; TP = temporal pole; FG = fusiform gyrus.

**Figure S2.**
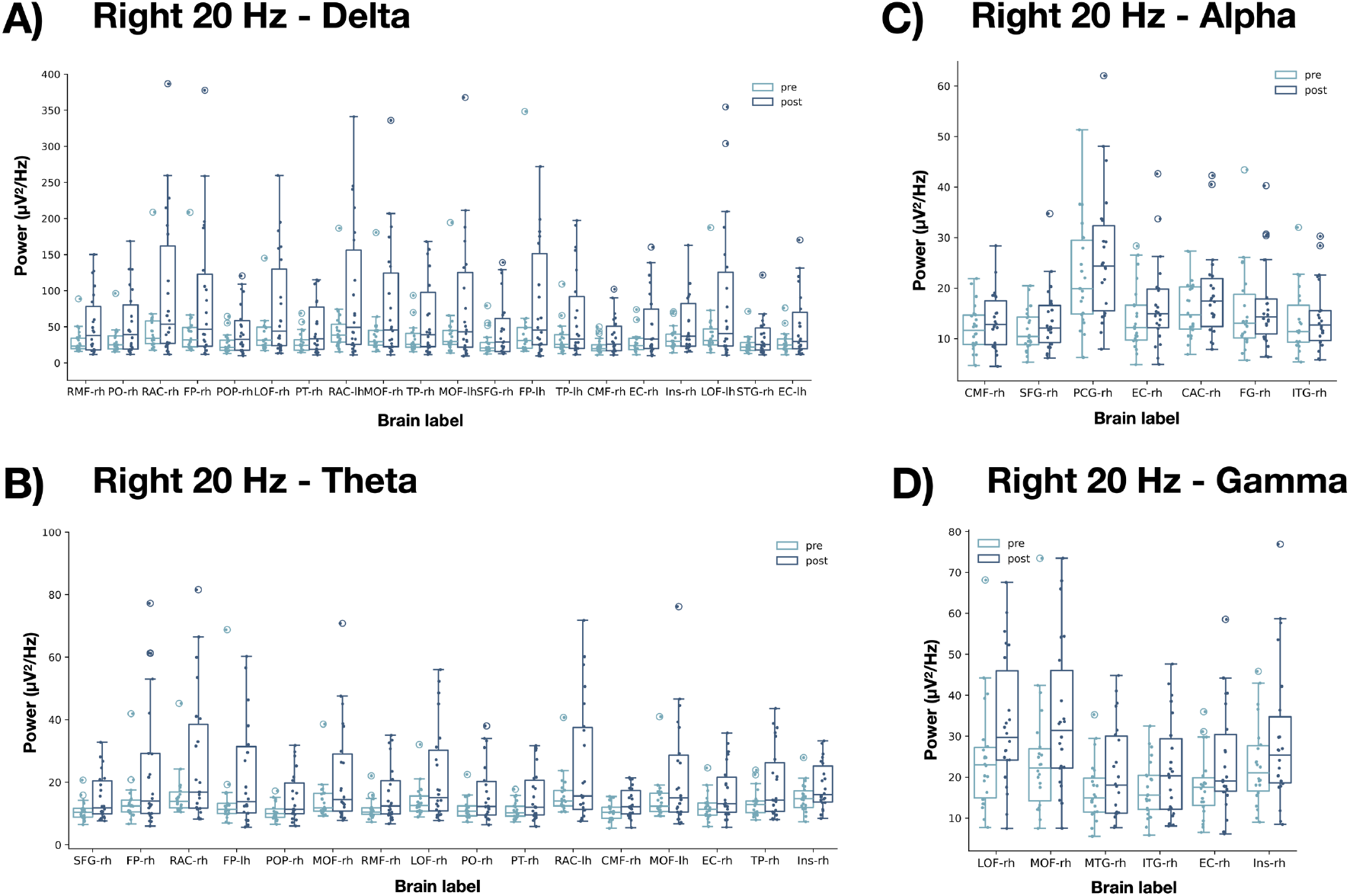
Pre and post EEG power values (µV^2^/Hz) for right 20 Hz rTMS. Boxplots illustrate pre and post stimulation EEG power values (µV^2^/Hz) for identified brain labels with a statistically significant and sham-superior power spectra increase in the **A)** Delta (0.5-4 Hz) and **B)** Theta (4-8 Hz) **C)** Alpha (8-13 Hz) and **D)** Gamma (30-80 Hz) frequency band following a stimulation of the right TPJ with 20 Hz rTMS. lh = left hemisphere; rh = right hemisphere; LOF = lateral orbitofrontal cortex; RAC = rostral anterior cingulate cortex; FP = frontal pole; MOF = medial orbitofrontal cortex; TP = temporal pole; EC = entorhinal cortex; STG = superior temporal gyrus; CMF = caudal middle frontal gyrus; POP = pars opercularis; PT = pars triangularis; RMF =rostal middle frontal gyrus; PO = pars orbitalis; Ins = Insula; SFG = superior frontal gyrus; ITG = inferior temporal gyrus; CAC = caudal anterior cingulate cortex; PCG = posterior cingulate cortex; FG = fusiform gyrus; MTG = middle temporal gyrus.

**Figure S3.**
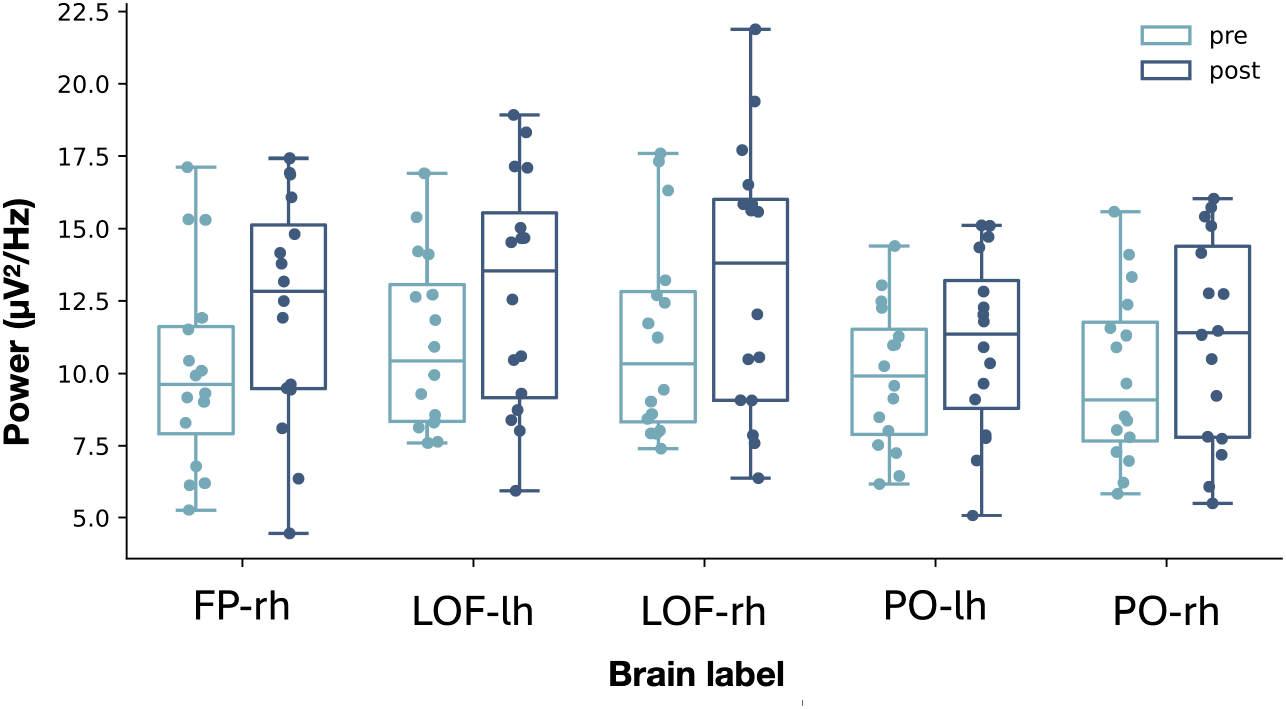
Pre and post EEG power values (µV^2^/Hz) for best protocols in the Alpha (8-13 Hz) frequency range. Boxplots illustrate pre and post stimulation EEG power values (µV^2^/Hz) in the Alpha (8-13 Hz) frequency band for identified brain labels following a stimulation with the individual best protocol for sham-superior and maximum tinnitus loudness reduction. lh = left hemisphere; rh = right hemisphere; FP = frontal pole, LOF = lateral orbitofrontal cortex, PO = pars orbitalis.

**Table S1:** Protocols with strongest and sham-superior tinnitus suppression per subject.

| Subject | Protocol | Average tinnitus suppression (%) |
| --- | --- | --- |
| S01 | 1Hz-rh | -17.14 |
| S02 | - | - |
| S03 | 10Hz-rh | -13.57 |
| S04 | 20Hz-rh | -39.29 |
| S05 | 10Hz-rh | -24.29 |
| S06 | 10Hz-lh | -10.00 |
| S07 | 1Hz-rh | -17.14 |
| S08 | 20Hz-lh | -7.86 |
| S09 | 1Hz-rh | -9.29 |
| S10 | 1Hz-lh | -26.43 |
| S11 | 10Hz-rh | -28.57 |
| S12 | 20Hz-lh | -24.29 |
| S13 | - | - |
| S14 | - | - |
| S15 | - | - |
| S16 | 1Hz-lh | -11.43 |
| S17 | - | - |
| S18 | - | - |
| S19 | 20Hz-lh | -14.29 |
| S20 | 1Hz-lh | -2.86 |
| S21 | 10Hz-rh | -12.86 |
| S22 | 20Hz-lh | -10.71 |
| All | 10Hz-rh: n = 4<br>20Hz-lh: n = 4<br>1Hz-lh: n = 3<br>1Hz-rh: n = 3<br>10Hz-lh: n = 1<br>20Hz-rh: n = 1 | -16.88 ± 9.40 |

lh = left hemisphere; rh = right hemisphere

